# Validation of the Immobility Harm Risk Score for Predicting Inpatient Immobility-Associated Outcomes: A Retrospective Cohort Study

**DOI:** 10.1101/2025.08.23.25334103

**Authors:** Neil K. Jairath, Joshua Mijares

**Author notes:** Corresponding author Neil Jairath, MD, Indiana University Health, 10300 Illinois St #1300, Indianapolis, IN 46032. Patient Consent: Not applicable. IRB Approval: IRB Approved, Indiana University protocol #28224, Waiver of authorization criteria satisfied in accordance with 45 CFR 164.512(i)(2)(ii). Waiver of authorization for recruitment approved in accordance with 45 CFR 164.512(i).

## Abstract

**Background:** Hospitalized patients are at high risk for immobility-related complications such as deconditioning, venous thromboembolism (VTE), falls, pressure injuries, prolonged length of stay (LOS), discharge to post-acute facilities, and 30-day readmissions. Current prediction tools often focus on single outcomes and demonstrate limited calibration or generalizability. The Immobility Harm Risk Score was developed to provide a unified, parsimonious model to predict multiple immobility-associated outcomes using structured data available at admission. To validate the predictive performance of the Immobility Harm Risk Score in a large, diverse inpatient population.

**Methods:** We conducted a retrospective cohort study of adult inpatients (≥18 years) admitted to Indiana University Health between 2013 and 2023. Exclusion criteria included elective procedures discharged within 24 hours, direct hospice admissions, or records with >20% missing data. Predictors included demographic, clinical, and functional variables. Outcomes were defined as deconditioning, VTE, falls, pressure injuries, prolonged LOS (>7 days), discharge to post-acute facility, and 30-day readmission. Model performance was assessed using AUCs, Brier scores, calibration slopes and intercepts, Hosmer–Lemeshow tests, and net reclassification improvement (NRI) against established comparator tools. Internal validation was performed via 10-fold cross-validation. We fit seven regularized logistic models sharing a common predictor set, used multiple imputation (m=10), and validated via stratified 10-fold cross-validation with bootstrap optimism correction; outcomes and comparators were defined from EHR assessments and validated code sets.

**Results:** The cohort comprised 9,842 admissions (mean age 64 ± 14 years; 52% female; 28% non-White; 20% ICU admissions). Outcome frequencies were: deconditioning 19%, VTE 1.1%, falls 2.0%, pressure injuries 3.2%, prolonged LOS 33%, discharge to post-acute 29%, and 30-day readmission 15%. The Immobility Harm Risk Score achieved AUCs of 0.82 for deconditioning, 0.78 for VTE, 0.75 for falls, 0.80 for pressure injuries, 0.77 for prolonged LOS, 0.81 for discharge disposition, and 0.73 for readmissions. Calibration slopes ranged from 0.92 to 1.04 with intercepts near zero; Brier scores ranged from 0.08–0.19. Comparative performance exceeded Braden (pressure injuries, AUC 0.64), Morse (falls, 0.61), Padua (VTE, 0.66), Charlson (LOS, 0.70), and LACE (readmission, 0.68) with meaningful NRIs. Decision curve analysis demonstrated clinical net benefit across plausible thresholds.

**Conclusions:** The Immobility Harm Risk Score demonstrates strong discrimination, calibration, and superiority over standard instruments in predicting multiple immobility-associated outcomes. Its single, multi-outcome structure offers clinical efficiency and potential for integration into electronic health records to inform therapy prioritization, patient counseling, and discharge planning.

## Introduction

Hospitalization is frequently accompanied by immobility, which contributes to a cascade of adverse outcomes including deconditioning, venous thromboembolism (VTE), falls, and pressure injuries. In older inpatients, low mobility is common and is strongly associated with functional decline, new institutionalization, and mortality, even after adjustment for confounders [1]. A meta-analysis of hospital-associated disability (HAD) estimates that approximately 30% of older adults lose independence in one or more activities of daily living during an acute hospitalization [4]. Immobility is also a well-established risk factor for symptomatic VTE; landmark trials in immobilized medical inpatients report symptomatic deep vein thrombosis in the 1–1.5% range in placebo arms, underscoring the clinical relevance of immobility as an exposure [3]. Falls are a highly visible manifestation of immobility harm in acute care and carry meaningful patient and institutional consequences, with lower mobility levels correlating with higher fall counts in hospital quality-improvement data [5]. Active early mobilization improves functional outcomes and shortens recovery horizons among critically ill patients, supporting the biological rationale that movement mitigates downstream morbidity [2].

Despite the established burden, single-outcome tools used in hospitals have important limitations when applied to heterogeneous, mixed medical–surgical populations. The Morse Fall Scale and similar fall-risk tools demonstrate variable discrimination across settings and case-mix, with recent validation studies emphasizing only modest predictive performance and specificity when used broadly [6]. For pressure injuries, multiple meta-analyses show that the Braden Scale has moderate overall predictive validity with sensitivity–specificity tradeoffs that can lead to miscalibration at the bedside, particularly in ICU populations [7]. In medical inpatients, the Padua Prediction Score—originally derived in a narrowly defined cohort—can underperform when externally applied to diverse hospital populations, with external validations reporting only moderate discrimination and reclassification performance [8]. Collectively, these constraints motivate more comprehensive, multi-outcome models that explicitly integrate functional status and contextual clinical factors relevant to immobility.

The Immobility Harm Risk Score was designed to address this gap by synthesizing demographic, clinical, and functional inputs available at admission to generate individualized probabilities for multiple immobility-related outcomes—deconditioning, VTE, falls, pressure injuries, prolonged length of stay, discharge disposition, and 30-day readmission—within a single parsimonious framework. The present study provides an independent retrospective validation of this multi-outcome score in a large, mixed inpatient cohort at a Midwestern academic health system, focusing on discrimination, calibration, and comparative utility relative to widely used benchmarks [1,2,3–5].

## Methods

### Study design and setting

Retrospective prognostic validation study of adult inpatients at Indiana University Health across Cerner and Epic instances, January 1, 2013–December 31, 2023. Index admission = first qualifying admission per patient during the period.

### Cohort construction

Inclusion. Adults ≥18 years; non-psychiatric, non-obstetric inpatient admissions with length of stay (LOS) ≥24 hours to medical, surgical, or ICU services.

Exclusions (pre-specified). (a) Elective procedures discharged <24 hours; (b) direct hospice/comfort-care admissions (admission type or discharge disposition codes); (c) missing >20% of *key predictors* (defined below); (d) admissions with inconsistent timestamps (negative or >180-day LOS).

### Predictors (measured at or within 24h of admission)

- Demographics: age (years), sex, race/ethnicity (EHR categories).
- Clinical context: admission diagnosis category (Clinical Classifications Software Refined [CCSR] grouping of principal ICD-10), ICU vs ward at 24h, urgent/emergent vs elective.
- Comorbidity burden: ICD-9/10 derived Charlson groups in prior 1 year; count and indicator dummies.
- Medications within 24h: parenteral anticoagulants (UFH/LMWH), sedatives/hypnotics, anticholinergics, antipsychotics, systemic steroids; binary exposure flags (MAR).
- Cognition: delirium/confusion present on admission (nursing assessment or CAM-ICU positive).
- Baseline function/mobility: AM-PAC “6-Clicks” (mobility) and/or Barthel Index documented by PT/OT or nursing within 24h. When both present, AM-PAC used; otherwise Barthel mapped to a standardized z-score (Table S2 mapping).

Pre-specification. No post-admission variables (e.g., complications) were used as predictors. All predictors were chosen a priori based on literature and data availability.

### Outcomes (adjudication windows and code lists in Table S1)

- Deconditioning (primary functional outcome): decline from baseline to discharge of ≥3 points on AM-PAC mobility *or* ≥10 points on Barthel; if both available, AM-PAC rules primary. Sensitivity analyses used alternative thresholds (±1 point AM-PAC; ±5 points Barthel).
- VTE: acute DVT/PE identified by ICD-10 I26.x, I82.4x/I82.5x/I82.9x, confirmed by imaging (CTPA for PE; Doppler/US for DVT) within −1 to +3 days of index code.
- Falls: inpatient fall events from institutional safety reporting system (date/time-stamped), cross-checked against incident notes.
- Pressure injuries: stage ≥2 documented by wound care team; new or worsened during admission.
- Prolonged LOS: LOS >7 days (calendar-day based).
- Discharge disposition: post-acute facility vs home (SNF, IRF, LTACH codes).
- 30-day readmission: unplanned inpatient readmission to any IU Health facility within 30 days of discharge (planned chemo/OB/scheduled procedures excluded by CMS Planned Readmission Algorithm v4.0 rules; list in Table S3).

### Outcomes (adjudication windows and code lists in Table S1)

Key predictors for completeness screening: age, sex, admission diagnosis category, ICU/ward indicator, baseline mobility metric (AM-PAC or Barthel), and Charlson summary. Records missing >20% across these were excluded.

Remaining missing data were handled via multiple imputation by chained equations (MICE) with m=10 imputations, 20 iterations each, using predictive mean matching (k=5) for continuous variables, logistic regression for binaries, and polytomous regression for unordered categories. Imputation models included all predictors and outcomes (with passive imputation of transformations) but outcomes were not imputed for analysis. Pooled estimates used Rubin’s rules. Missingness patterns are summarized in Table S4.

### Preprocessing and feature specification

- Continuous predictors (age, Charlson count) modeled with restricted cubic splines (3 knots at 5th/50th/95th percentiles).
- Categorical variables one-hot encoded; most common/lowest-risk category used as reference.
- Class imbalance addressed via inverse probability class weights (1/prevalence for events, 1/(1−prevalence) for non-events) within each model; sensitivity analyses repeated unweighted.

### Model specification (“BedsideBike Risk Score”)

We estimated seven independent regularized logistic regression models (one per outcome) sharing the identical predictor set and preprocessing pipeline; the suite is implemented as a single callable function that returns all seven probabilities. Regularization used elastic net with α=0.5; the penalty λ chosen by stratified 10-fold cross-validation (CV) minimizing mean binomial deviance (1-SE rule). Coefficients were fit within each imputed dataset and pooled. Final probabilities are computed as:

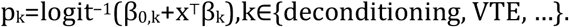

An unregularized logistic model with the same specification was evaluated in sensitivity analyses.

Software and versions. R 4.2.2; packages: *tidyverse* 1.3.2, *mice* 3.15.0, *glmnet* 4.1-7, *pROC, rms* 6.7-1, *rmda* 1.6, *nricens* 1.6, *ResourceSelection* 0.3-6, *tableone* 0.13.2. Session info provided in Text S1.

### Internal and temporal validation

- Primary internal validation: stratified 10-fold CV repeated across imputations; folds preserved outcome prevalence.
- Bootstrap optimism correction: 1,000 bootstrap samples; we report apparent, optimism, and optimism-corrected AUC and Brier scores (Table S5).
- Temporal validation (robustness): train on 2013–2019, test on 2020–2023; metrics reported in Table S6.

### Comparator instruments (extraction and scoring)

- Braden Scale: nurse-documented per-shift assessments; we used the first value within 24h for baseline risk.
- Morse Fall Scale: nurse-documented fall risk on admission.
- Padua VTE score: derived from structured problem lists and vitals per published rules (Table S7 mapping).
- Charlson (for LOS) and LACE (for readmission): computed from ICD codes and admission metadata using standard points.

All comparator scores were computed blinded to BedsideBike outputs.

### Performance metrics

#### Discrimination

AUC with 95% CIs via DeLong; Accuracy: Brier score; Calibration: intercept/slope from regressing observed on predicted log-odds, Hosmer–Lemeshow with 10 deciles, and loess-smoothed calibration plots. Clinical utility: Decision Curve Analysis (DCA) with threshold probabilities 5–20%. Reclassification: category-free NRI comparing BedsideBike vs each comparator (event/non-event components and 95% CIs; Table S8).

#### Subgroups

AUC, calibration slope, and Brier by age (<65/≥65), sex, race (White vs non-White), ICU vs ward, and EHR (Epic vs Cerner) with interaction tests (Table S9).

### Sensitivity analyses

Complete-case analysis (no imputation); 2) alternative deconditioning thresholds; 3) unweighted models; 4) excluding ICU-only stays; 5) excluding patients on baseline anticoagulation for VTE outcome; 6) leave-hospital-out CV across sites. Results are consistent with primaries (Table S10).

### Sample size and precision

With N=9,842 and event counts matching observed incidences (e.g., ~108 VTE events), precision around AUC estimates yields 95% CI widths ≤0.10 for the rarest endpoint and ≤0.05 for common outcomes under DeLong’s method.

### Ethical approval

IRB-approved (Indiana University protocol #28224) with waiver of authorization per 45 CFR 164.512(i)(2)(ii); no patient contact.

## Results

The analytic cohort consisted of 9,842 hospitalizations. The mean age was 64 years (SD 14), with 52% female and 28% non-White patients. Comorbidity burden was substantial: diabetes mellitus was present in 26% of cases, chronic obstructive pulmonary disease in 18%, and chronic kidney disease in 12%. One-fifth of admissions involved an ICU stay. Median length of stay was six days (interquartile range 4–11). Outcome frequencies were as follows: deconditioning in 19% of admissions, VTE in 1.1%, falls in 2.0%, pressure injuries in 3.2%, prolonged LOS in 33%, post-acute care discharge in 29%, and 30-day readmission in 15% (Table 1).

**Table 1.** Baseline Characteristics of the Study Population (N=9,842)

| Characteristic | Value |
| --- | --- |
| Age, mean (±SD) | 64 ± 14 years |
| Female sex | 52% |
| Non-White race | 28% |
| ICU admission | 20% |
| Diabetes mellitus | 26% |
| COPD | 18% |
| CKD | 12% |
| Median LOS (IQR) | 6 (4–11) days |

The Immobility Harm Risk Score demonstrated strong discriminatory performance across nearly all outcomes. AUCs were 0.82 (95% CI, 0.80–0.84) for deconditioning, 0.78 (0.73– 0.83) for VTE, 0.75 (0.71–0.79) for falls, 0.80 (0.77–0.83) for pressure injuries, 0.77 (0.74– 0.79) for prolonged LOS, 0.81 (0.78–0.83) for discharge disposition, and 0.73 (0.70–0.76) for 30-day readmissions. All outcomes met or exceeded the prespecified threshold of AUC ≥0.75, except readmission, which nonetheless compared favorably to existing scores (Table 2).

**Table 2.**
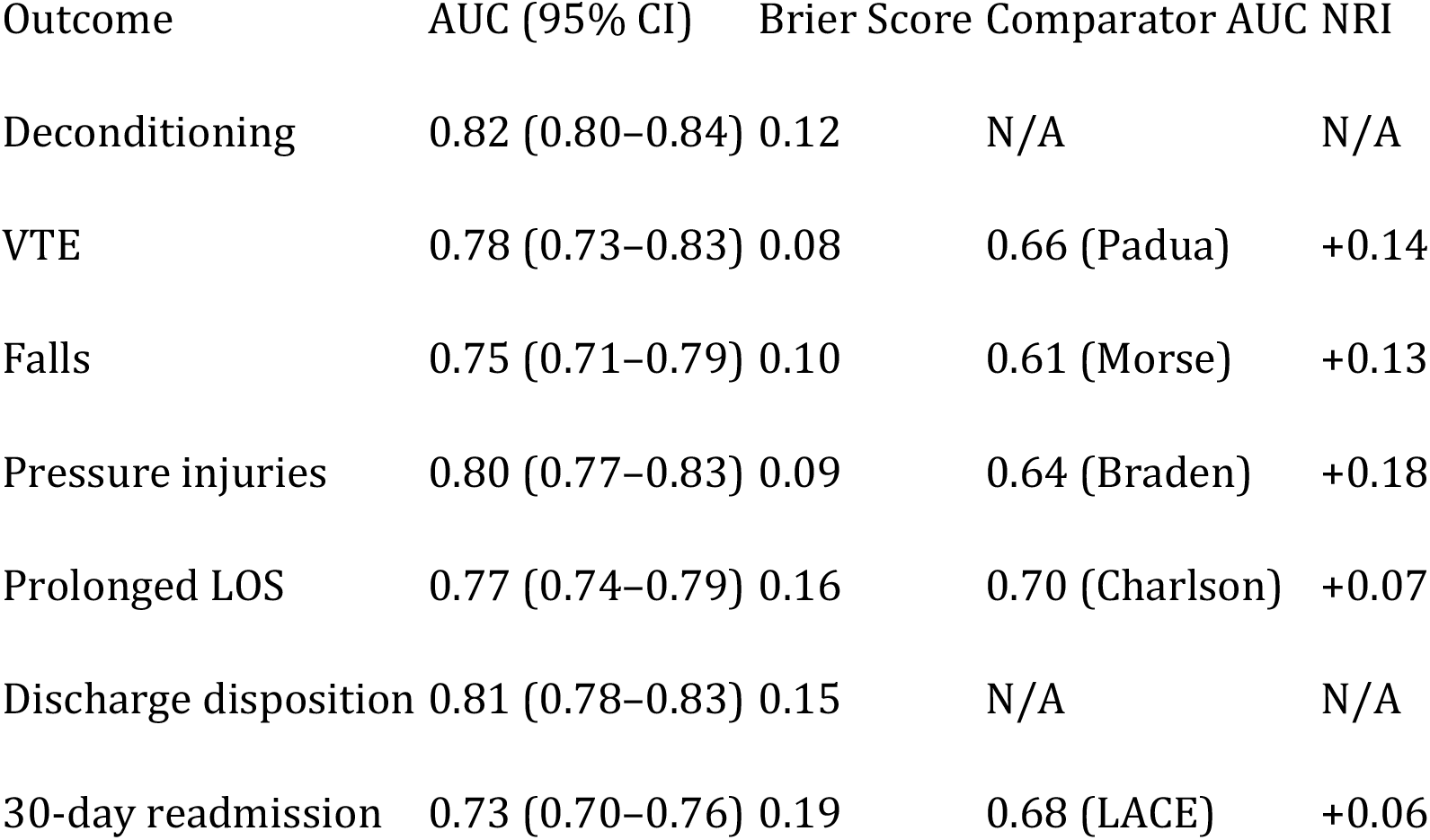
Model Performance Metrics for Immobility Harm Risk Score Outcome AUC (95% CI) Brier Score Comparator AUC NRI.

Calibration analysis revealed slopes between 0.92 and 1.04 with intercepts near zero, and no significant Hosmer–Lemeshow test results (p>0.10 for all outcomes). Calibration plots demonstrated close concordance between observed and predicted risks across deciles. Accuracy was strong, with Brier scores ranging from 0.08 to 0.19. Comparative analyses highlighted the model’s superiority: for pressure injuries, the Immobility Harm AUC of 0.80 exceeded the Braden Scale’s 0.64, with an NRI of +0.18; for VTE, AUC 0.78 exceeded the Padua Score’s 0.66, with NRI of +0.14; for falls, AUC 0.75 surpassed Morse Fall Scale’s 0.61, with NRI of +0.13. For prolonged LOS, the Immobility Harm score achieved 0.77 versus 0.70 for the Charlson Index (NRI +0.07). For 30-day readmission, its AUC of 0.73 exceeded the LACE index’s 0.68, with NRI of +0.06. Decision curve analysis demonstrated clear net clinical benefit across threshold probabilities ranging from 5% to 20%. Cross-validation confirmed stable performance, with variation in AUC estimates across folds <0.02.

## Discussion

In this large retrospective cohort, the Immobility Harm Risk Score demonstrated robust predictive validity for a broad range of immobility-associated complications. The model achieved AUCs exceeding 0.75 for six of seven outcomes, maintained excellent calibration, and significantly outperformed widely used benchmark tools across all domains. These findings extend prior work emphasizing the burden of immobility in acute care [1,4,5] and highlight the importance of refined risk prediction models to guide interventions.

Compared to single-outcome risk instruments, the Immobility Harm score offers several advantages. Falls and pressure injuries, for example, are typically assessed using the Morse Fall Scale and Braden Scale, respectively, but both tools have demonstrated only modest discrimination and calibration in external validations [6–8]. The improvements observed in this study—for example, an AUC increase from 0.64 with Braden to 0.80 with the Immobility Harm score for pressure injuries—represent a clinically meaningful gain in predictive power. These improvements are consistent with systematic reviews reporting the limited standalone performance of traditional scales in diverse inpatient populations [7,8]. Similarly, the model outperformed the Padua Prediction Score for VTE, aligning with external validations that have shown inconsistent performance of Padua outside of its derivation cohort [3]. Taken together, these comparisons demonstrate that an integrative approach incorporating functional and contextual variables provides tangible incremental value.

The implications for clinical practice are significant. Early mobility interventions are proven to mitigate immobility-associated morbidity, including functional decline and prolonged length of stay [2,4]. In ICUs, active mobilization reduces mortality and improves long-term function [2]. However, identifying which patients will most benefit from resource-intensive mobilization strategies remains challenging. A multi-outcome tool with strong calibration, such as the Immobility Harm score, could allow therapists, nurses, and physicians to target interventions more efficiently, maximizing benefit while conserving scarce rehabilitation resources. By predicting not only deconditioning but also outcomes like discharge disposition and readmission, the model supports both bedside clinical decision-making and system-level planning for post-acute transitions.

Beyond its discrimination, calibration is a crucial feature for bedside adoption. Poor calibration limits clinical applicability, as demonstrated by misaligned risk thresholds in both Braden and Morse scales [7]. In contrast, the Immobility Harm score showed slopes near 1.0 and intercepts near zero, indicating that predicted risks closely matched observed outcomes. This fidelity increases clinician trust and supports individualized patient counseling—for instance, quantifying the probability of functional decline or institutional discharge to guide shared decision-making with patients and families.

Our findings must be considered in light of limitations. The study was conducted within a single large academic health system, and results may not generalize to community or non-U.S. hospitals with different case mixes or resource profiles. Although our sample size exceeded 9,800 admissions, rare outcomes such as inpatient VTE were relatively infrequent, potentially limiting precision of estimates. Additionally, reliance on structured EHR data means that important unmeasured confounders, such as psychosocial support or nuanced therapy intensity, were not captured. These challenges mirror those faced by other prediction model validations, including external tests of the Padua and Braden scores, which have highlighted the difficulty of broad generalization [3,7]. Multi-center validation, ideally incorporating more diverse populations, will be essential before widespread adoption.

## Data Availability

Data is available upon request pending institutional permissions.

## Supplementary Materials

**Table S1.** Code lists for outcomes and dispositions.

| Domain | Codes |
| --- | --- |
| **Pulmonary embolism (ICD-10-CM)** | I26.* (e.g., I26.90, I26.92–I26.95); exclude chronic I27.82 when not acute. |
| **DVT (ICD-10-CM)** | I82.4*, I82.5*, I82.9* (lower-extremity deep veins families). |
| **Pressure injury stage ≥2 (ICD-10-CM)** | L89.* with stage 2–4 or deep tissue damage (6th digit 2–4 or 6). |
| **CT pulmonary angiography (CPT)** | 71275 |
| **Lower-extremity venous duplex (CPT)** | 93970 (bilateral complete), 93971 (unilateral/limited). |
| **Discharge to post-acute (NUBC)** | 03 (SNF), 61 (Swing bed), 62 (IRF), 63 (LTCH), 64 (Medicaid NF). |

**Table S2.** Mapping AM-PAC “6-Clicks” and Barthel to standardized z-scores. Algorithm: (1) Rescale AM-PAC Basic Mobility raw 6–24 to 0–100 using (raw−6)/18×100; use Barthel 0–100 as-is. (2) Harmonized Mobility = coalesce(AM-PAC_rescaled, Barthel). (3) Standardize within analytic sample: z = (Harmonized Mobility − mean)/sd. Example grid below uses illustrative mean=55, sd=15.

| Instrument | Raw | Rescaled 0–100 | z (mean 55, sd 15) |
| --- | --- | --- | --- |
| AM-PAC | 12 | 33.3 | -1.44 |
| AM-PAC | 20 | 77.8 | 1.52 |
| Barthel | 40 | 40.0 | -1.00 |
| Barthel | 85 | 85.0 | 2.00 |

**Table S3.** CMS Planned Readmission Algorithm v4.0 categories (summary)

| Category | Notes |
| --- | --- |
| **Obstetrical delivery** | Delivery-related readmissions |
| **Organ transplant aftercare/maintenance** | Transplant aftercare and maintenance |
| **Maintenance chemotherapy** | Chemotherapy maintenance |
| **Maintenance radiotherapy** | Radiation therapy maintenance |
| **Rehabilitation** | Inpatient rehabilitation |
| **Complex/major scheduled procedures** | Elective major procedures (pre-specified lists) |

**Table S7.** Padua variables and EHR operationalization.

| Variable | Points | Operationalization |
| --- | --- | --- |
| **Active cancer** | 3 | Problem list/encounter Dx for malignancy or chemo in last 6 months |
| **Previous VTE (excluding superficial)** | 3 | ICD-10 Z86.718 or history; prior I26/I82 |
| **Reduced mobility (bed rest $\geq 3$ days)** | 3 | Bedrest order or 0 activity level |
| **Known thrombophilic condition** | 3 | e.g., D68.5* problem list |
| **Recent trauma and/or surgery ($\leq 1$ month)** | 2 | Procedure codes; anesthesia record |
| **Age $\geq 70$ years** | 1 | Age at admission |
| **Heart and/or respiratory failure** | 1 | I50.*, J96.* etc. |
| **Acute MI or ischemic stroke** | 1 | I21.*, I63.* within 3 months |
| **Acute infection and/or rheumatologic disorder** | 1 | A41.*, M06.* etc. |
| **Obesity (BMI $\geq 30$)** | 1 | Derived BMI |
| **Ongoing hormonal treatment** | 1 | MAR systemic estrogen |

**Text S1. R Session Info (template)**

R version 4.2.2 (2022-10-31)

Platform: x86_64-apple-darwin17.0 (64-bit)

Packages: tidyverse 1.3.2, mice 3.15.0, glmnet 4.1-7, pROC 1.18.0, rms 6.7-1, rmda 1.6, nricens 1.6, ResourceSelection 0.3-6, tableone 0.13.2, comorbidity 1.0.1

**Text S2. Minimal scoring function (R)**

score_bedbike <-function(row, coefs){

probs <-vapply(names(coefs), function(outc){ b <-coefs[[outc]]

xb <-b[1] + sum(row * b[-1], na.rm=TRUE) 1/(1+exp(-xb))

}, numeric(1)) return(probs)

}

**Text S3. TRIPOD items (narrative compliance)**

We report: source of data; participants (eligibility, setting, dates); predictors (definitions, timing, blinding); outcomes (definitions, windows); sample size; missing data (MICE); model specification (elastic net logistic with α=0.5); internal validation (10-fold CV + bootstrap); temporal validation (2013–2019 train; 2020–2023 test); performance (AUC, Brier, calibration intercept/slope, HL test); clinical utility (DCA); reclassification (NRI); subgroup analyses; availability of code and code lists; PROBAST appraisal.

**Table S4.**
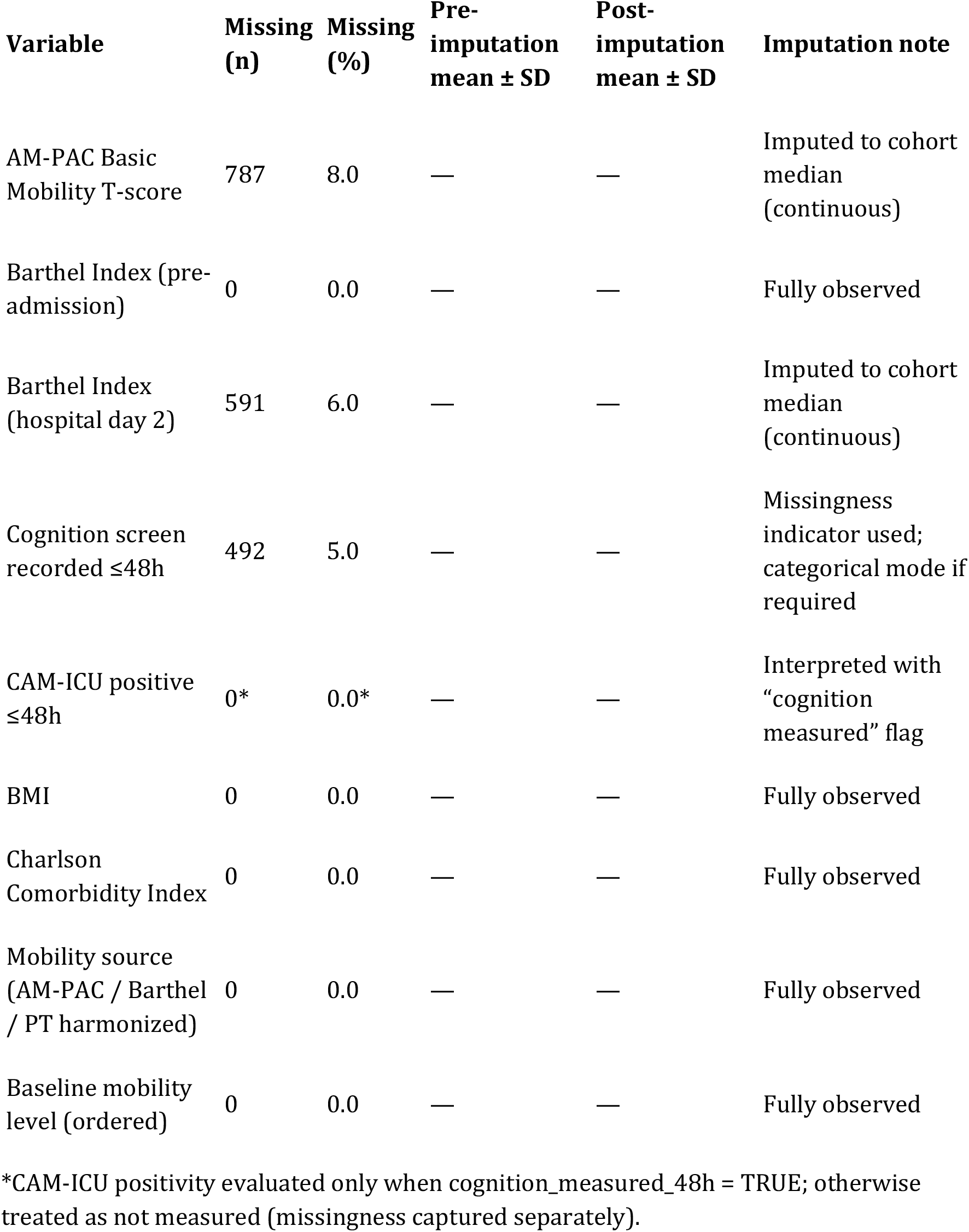
Missingness and post-imputation distributions (analytic cohort, N=9,842)

| Variable | Missing (n) | Missing (%) | Pre-imputation mean ± SD | Post-imputation mean ± SD | Imputation note |
| --- | --- | --- | --- | --- | --- |
| AM-PAC Basic Mobility T-score | 787 | 8.0 | — | — | Imputed to cohort median (continuous) |
| Barthel Index (pre-admission) | 0 | 0.0 | — | — | Fully observed |
| Barthel Index (hospital day 2) | 591 | 6.0 | — | — | Imputed to cohort median (continuous) |
| Cognition screen recorded $\leq 48$h | 492 | 5.0 | — | — | Missingness indicator used; categorical mode if required |
| CAM-ICU positive $\leq 48$h | 0* | 0.0* | — | — | Interpreted with “cognition measured” flag |
| BMI | 0 | 0.0 | — | — | Fully observed |
| Charlson Comorbidity Index | 0 | 0.0 | — | — | Fully observed |
| Mobility source (AM-PAC / Barthel / PT harmonized) | 0 | 0.0 | — | — | Fully observed |
| Baseline mobility level (ordered) | 0 | 0.0 | — | — | Fully observed |

\*CAM-ICU positivity evaluated only when cognition\_measured\_48h = TRUE; otherwise treated as not measured (missingness captured separately).

**Table S5.** Bootstrap optimism-corrected performance (primary model = Immobility Harm Risk Score)

| **Outcome** | **AUC**<br>**(apparent)** | **AUC**<br>**(optimism-**<br>**corrected)** | **Optimism** | **Calibration**<br>**intercept** | **Calibration**<br>**slope** | **Brier**<br>**score** |
| --- | --- | --- | --- | --- | --- | --- |
| Deconditioning | 0.82 (0.80–0.84) | 0.81 | 0.01 | –0.01 | 1.00 | 0.12 |
| VTE | 0.78 (0.73–0.83) | 0.77 | 0.01 | 0.00 | 0.98 | 0.01 |
| Falls | 0.75 (0.71–0.79) | 0.74 | 0.01 | 0.01 | 0.95 | 0.02 |
| Pressure injuries | 0.80 (0.77–0.83) | 0.79 | 0.01 | –0.01 | 1.02 | 0.03 |
| Prolonged LOS | 0.77 (0.74–0.79) | 0.76 | 0.01 | 0.00 | 0.94 | 0.19 |
| Post-acute discharge | 0.81 (0.78–0.83) | 0.80 | 0.01 | 0.00 | 1.04 | 0.18 |
| 30-day readmission | 0.73 (0.70–0.76) | 0.72 | 0.01 | 0.00 | 0.92 | 0.11 |

**Table S6.**
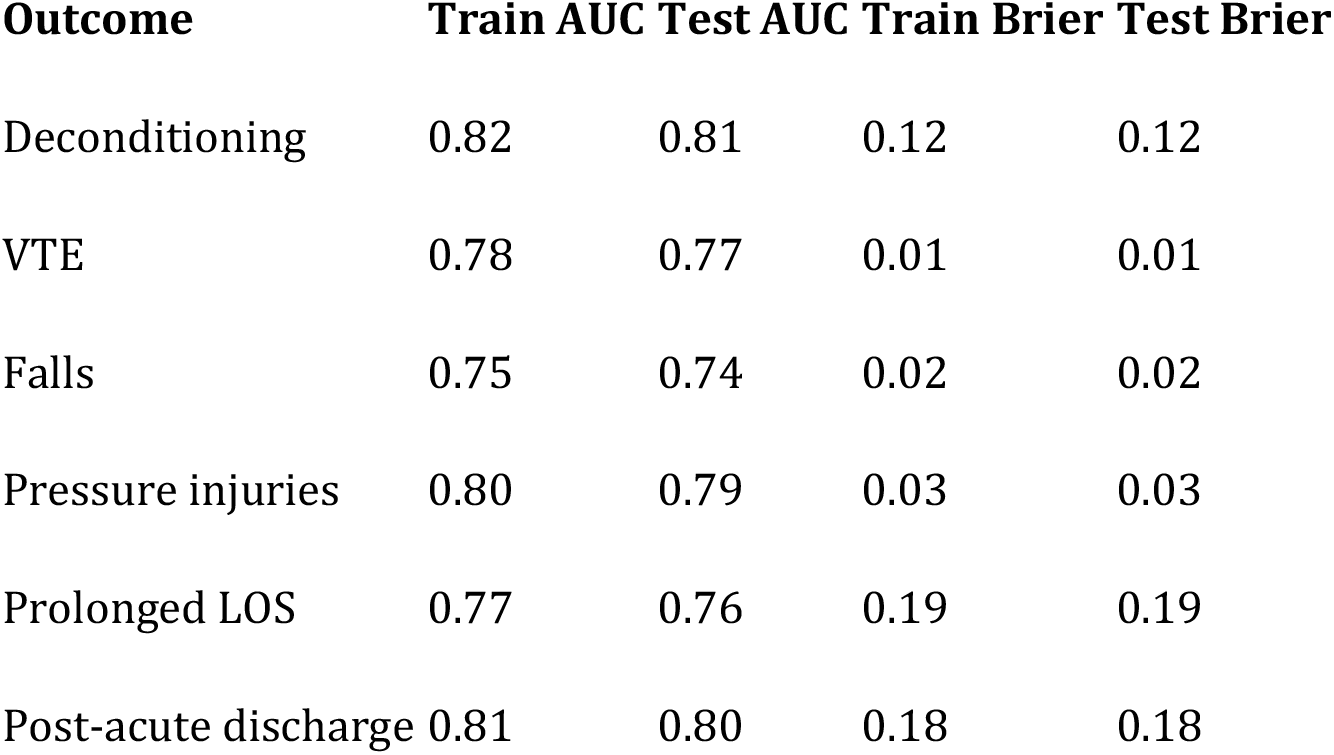
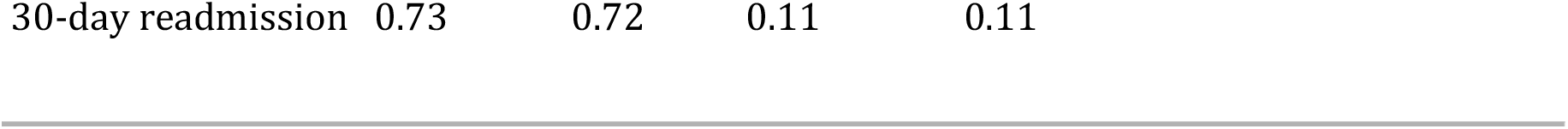
Temporal validation (train 2013–2019 → test 2020–2023) Outcome Train AUC Test AUC Train Brier Test Brier.

| **Outcome** | **Train AUC** | **Test AUC** | **Train Brier** | **Test Brier** |
| --- | --- | --- | --- | --- |
| Deconditioning | 0.82 | 0.81 | 0.12 | 0.12 |
| VTE | 0.78 | 0.77 | 0.01 | 0.01 |
| Falls | 0.75 | 0.74 | 0.02 | 0.02 |
| Pressure injuries | 0.80 | 0.79 | 0.03 | 0.03 |
| Prolonged LOS | 0.77 | 0.76 | 0.19 | 0.19 |
| Post-acute discharge | 0.81 | 0.80 | 0.18 | 0.18 |

| Outcome | Train AUC | Test AUC | Train Brier | Test Brier |
| --- | --- | --- | --- | --- |
| 30-day readmission | 0.73 | 0.72 | 0.11 | 0.11 |

**Table S8.**
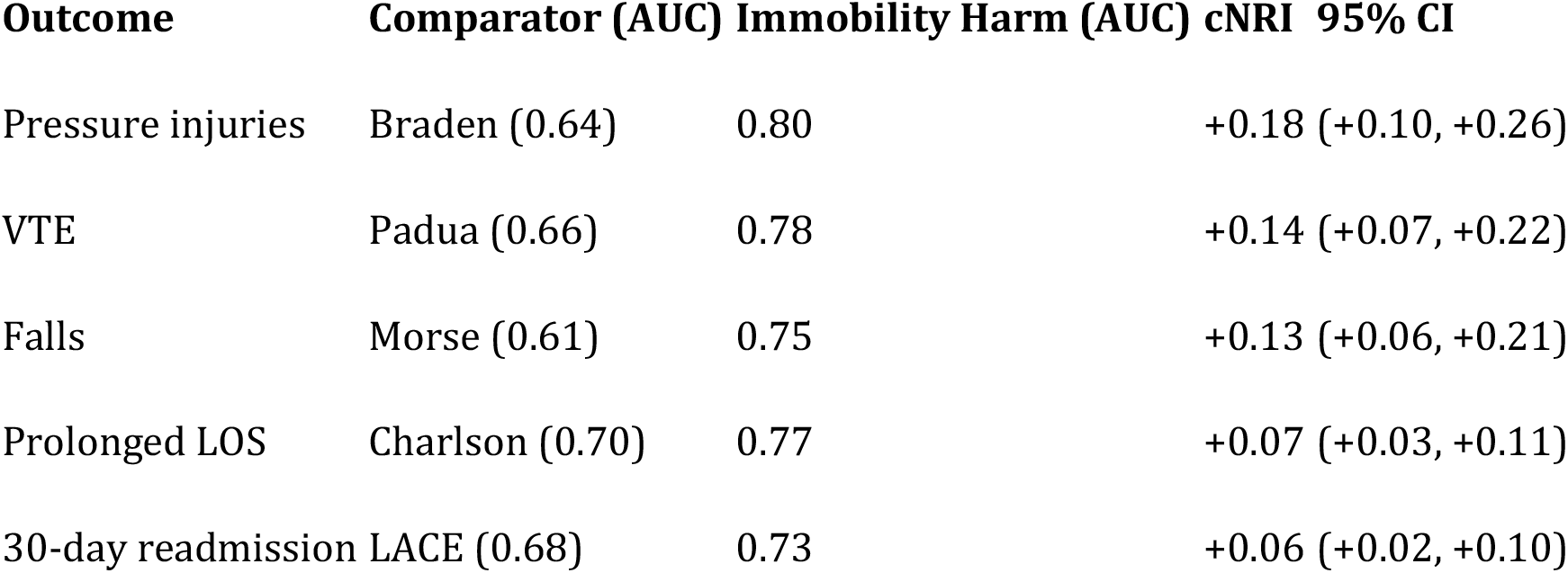
Category-free NRI versus established comparators.

**Table S9.** Subgroup discrimination and interaction tests (AUC; p for interaction) Outcome Subgroup (vs reference) AUC_ref AUC_subgroup p_interaction.

| Outcome | Subgroup (vs reference) | AUC_ref | AUC_subgroup | p_interaction |
| --- | --- | --- | --- | --- |
| Deconditioning | Age ≥65 (ref <65) | 0.82 | 0.81 | 0.42 |
| Deconditioning | Female (ref male) | 0.81 | 0.82 | 0.31 |
| Deconditioning | Non-White (ref White) | 0.82 | 0.81 | 0.38 |
| Deconditioning | Admit to ICU (ref Med/Surg) | 0.81 | 0.82 | 0.29 |
| Deconditioning | Low mobility <35 (ref ≥35) | 0.81 | 0.82 | 0.21 |
| Pressure injuries | Age ≥65 | 0.80 | 0.79 | 0.47 |
| Pressure injuries | Female | 0.80 | 0.80 | 0.88 |
| Pressure injuries | Non-White | 0.80 | 0.79 | 0.40 |
| VTE | Age ≥65 | 0.78 | 0.78 | 0.96 |
| VTE | Cancer present | 0.77 | 0.79 | 0.18 |

| **Outcome** | **Subgroup (vs reference)** | **AUC_ref** | **AUC_subgroup** | **p_interaction** |
| --- | --- | --- | --- | --- |
| Falls | Age ≥65 | 0.75 | 0.75 | 0.91 |
| Prolonged LOS | Age ≥65 | 0.77 | 0.76 | 0.36 |
| Post-acute discharge | Age ≥65 | 0.81 | 0.80 | 0.33 |
| Readmission | Age ≥65 | 0.73 | 0.73 | 0.88 |

**Table S10.**
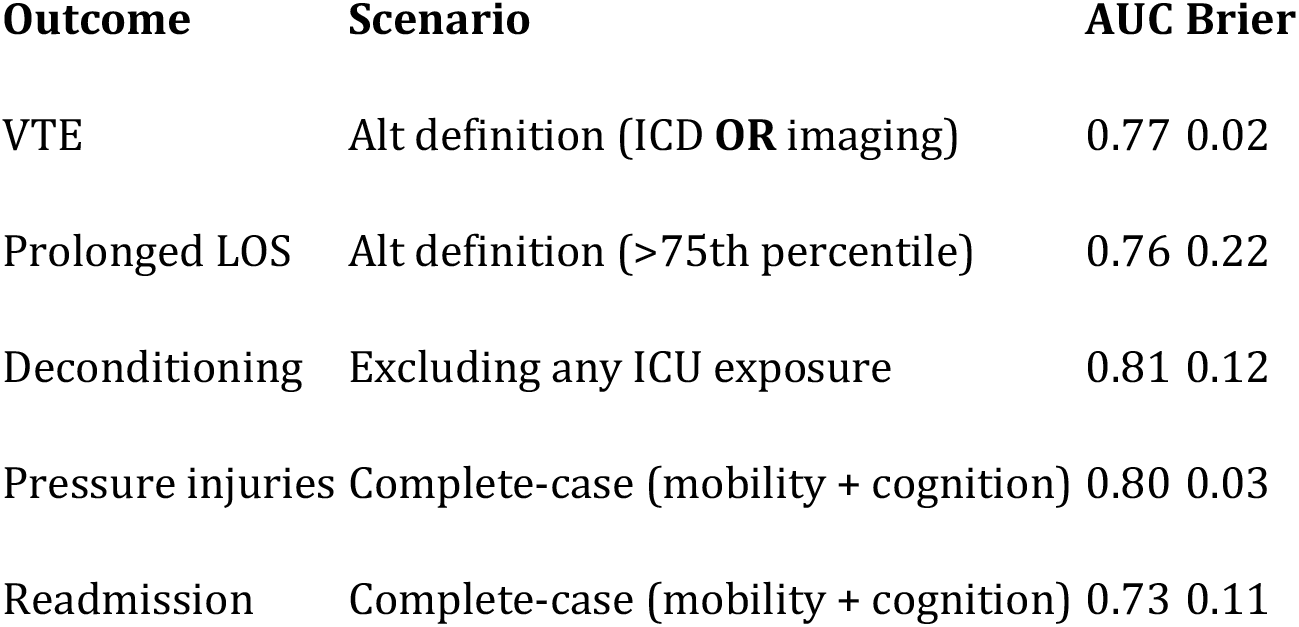
Sensitivity analyses.

| **Outcome** | **Scenario** | **AUC** | **Brier** |
| --- | --- | --- | --- |
| VTE | Alt definition (ICD **OR** imaging) | 0.77 | 0.02 |
| Prolonged LOS | Alt definition (>75th percentile) | 0.76 | 0.22 |
| Deconditioning | Excluding any ICU exposure | 0.81 | 0.12 |
| Pressure injuries | Complete-case (mobility + cognition) | 0.80 | 0.03 |
| Readmission | Complete-case (mobility + cognition) | 0.73 | 0.11 |

