## Supplementary material for "Validation of the Immobility Harm Risk Score for Predicting Inpatient Immobility-Associated Outcomes: A Retrospective Cohort Study": R code: 00_setup.R

```
# 00_setup.R — packages & options
required <- c(
  "tidyverse","data.table","janitor","mice","glmnet","pROC","rms","rmda",
  "nricens","ResourceSelection","tableone","comorbidity","flextable","officer","ggplot2"
)
ip <- installed.packages()[, "Package"]
to_install <- setdiff(required, ip)
if (length(to_install)) install.packages(to_install, repos="https://cloud.r-project.org")

invisible(lapply(required, library, character.only=TRUE))
options(stringsAsFactors = FALSE, scipen = 999)

# Set seed for reproducibility
set.seed(42)
```
