## Supplementary material for "Validation of the Immobility Harm Risk Score for Predicting Inpatient Immobility-Associated Outcomes: A Retrospective Cohort Study": R code: 01_data_spec.R

```
# 01_data_spec.R — expected input schemas and helper readers

# Expected data:
# admissions: id, patient_id, admit_time, discharge_time, los_hours, service, admission_class, admission_type, discharge_disposition, ehr ("Epic"|"Cerner")
# patients: patient_id, birth_date, sex, race_ethnicity
# units: admission_id, unit_type, time_from_admit_hours
# dx: admission_id, icd10_code, dx_type ("principal"|"secondary"), dx_date
# meds: admission_id, med_name, route, start_time, stop_time
# nursing: admission_id, cam_icu_positive (0/1), braden_total, morse_total, ampac_mobility_raw, barthel_total, assessment_time
# imaging: admission_id, cpt_code, order_time
# safety_events: admission_id, event_time, event_type ("fall"), severity

# Readers (replace with your DB extracts)
read_all <- function(path) {
  list(
    admissions = readr::read_csv(file.path(path, "admissions.csv")),
    patients   = readr::read_csv(file.path(path, "patients.csv")),
    units      = readr::read_csv(file.path(path, "units.csv")),
    dx         = readr::read_csv(file.path(path, "dx.csv")),
    meds       = readr::read_csv(file.path(path, "meds.csv")),
    nursing    = readr::read_csv(file.path(path, "nursing.csv")),
    imaging    = readr::read_csv(file.path(path, "imaging.csv")),
    safety     = readr::read_csv(file.path(path, "safety_events.csv"))
  )
}
```
