## Supplementary material for "Validation of the Immobility Harm Risk Score for Predicting Inpatient Immobility-Associated Outcomes: A Retrospective Cohort Study": R code: 02_code_lists.R

```
# 02_code_lists.R — ICD/CPT & rules

# Venous thromboembolism (ICD-10-CM)
icd10_pe <- c(
  # Pulmonary embolism w/ & w/o cor pulmonale and specified subtypes
  paste0("I26.", c("0","00","01","02","03","09","90","92","93","94","95","99"))
)
# Include general I26 pattern as safeguard:
icd10_pe_pattern <- "^I26"

icd10_dvt <- c(
  # Lower-extremity DVT families
  "I82.4", # acute embolism/thrombosis of deep veins of lower extremity (family)
  "I82.5", # embolism/thrombosis of iliac/femoral/popliteal etc. (family)
  "I82.9"  # embolism/thrombosis of unspecified vein
)
icd10_dvt_pattern <- "^I82\\.(4|5|9)"

# Pressure injury (ICD-10-CM L89* by site & stage >= 2 and deep tissue damage)
icd10_pressure_ulcer_pattern <- "^L89\\."

# CPT imaging confirmation
cpt_ctpa <- "71275"  # CT pulmonary angiography
cpt_venous_duplex <- c("93970","93971")  # LE venous duplex

# Discharge disposition (NUBC UB-04 patient status codes)
dispo_home <- c("01")
dispo_post_acute <- c("03","61","62","63","64") # SNF, swing bed, IRF, LTCH, Medicaid NF

# CMS Planned Readmission Algorithm v4.0 categories (diagnostic/ procedure groups)
cms_planned_categories <- c(
  "Obstetrical delivery","Organ transplant aftercare/maintenance",
  "Maintenance chemotherapy","Maintenance radiotherapy",
  "Rehabilitation","Complex/major scheduled procedures"
)

# Padua Prediction Score variables and points per Barbar et al. 2010 (JTH)
padua_vars <- tibble::tribble(
  ~variable, ~points, ~operationalization,
  "Active cancer", 3, "Problem list/encounter Dx for malignancy or chemo in last 6 months",
  "Previous VTE (excluding superficial)", 3, "ICD-10 Z86.718 or history; prior I26/I82",
  "Reduced mobility (bed rest ≥3 days)", 3, "Bedrest order or 0 activity level",
  "Known thrombophilic condition", 3, "E.g., D68.5*; problem list",
  "Recent trauma and/or surgery (≤1 month)", 2, "Procedure codes; anesthesia record",
  "Elderly age (≥70 years)", 1, "Age at admission",
  "Heart and/or respiratory failure", 1, "I50.*, J96.*, etc.",
  "Acute MI or ischemic stroke", 1, "I21.*, I63.* within 3 months",
  "Acute infection and/or rheumatologic disorder", 1, "A41.*, M06.*, etc.",
  "Obesity (BMI ≥30)", 1, "Height/weight derived BMI",
  "Ongoing hormonal treatment", 1, "MAR: systemic estrogen; gender-affirming as applicable"
)
```
