## Supplementary material for "Validation of the Immobility Harm Risk Score for Predicting Inpatient Immobility-Associated Outcomes: A Retrospective Cohort Study": R code: 03_extract_transform.R

```
# 03_extract_transform.R — cohort & feature derivation
source("R/02_code_lists.R")

construct_cohort <- function(d) {
  a <- d$admissions %>%
    mutate(age = as.integer(floor(as.numeric(difftime(admit_time, d$patients$birth_date[match(patient_id, d$patients$patient_id)], units="days"))/365.25))) %>%
    filter(age >= 18,
           los_hours >= 24,
           admission_class %in% c("Inpatient","Inpatient-ICU"),
           !service %in% c("Psychiatry","Obstetrics"))
  
  # Exclusion: elective <24h handled above; hospice/comfort care
  a <- a %>% filter(!discharge_disposition %in% c("50","51")) # hospice home/facility
  
  # Deduplicate to first qualifying admission per patient
  a <- a %>% arrange(patient_id, admit_time) %>%
    group_by(patient_id) %>% slice(1) %>% ungroup()
  
  a
}

derive_predictors <- function(d, cohort){
  idv <- cohort %>% select(admission_id = id, patient_id, admit_time, ehr = ehr)
  
  # Demographics
  demo <- d$patients %>% select(patient_id, sex, race_ethnicity)
  idv <- idv %>% left_join(demo, by="patient_id")
  idv$age <- cohort$age
  
  # Admission diagnosis category (CCSR placeholder: use principal ICD-10 to group)
  principal_dx <- d$dx %>% filter(dx_type=="principal") %>%
    arrange(admission_id, dx_date) %>% group_by(admission_id) %>% slice(1) %>% ungroup() %>%
    select(admission_id, icd10_code)
  idv <- idv %>% left_join(principal_dx, by="admission_id") %>% rename(principal_icd10 = icd10_code)
  
  # ICU vs ward at 24h
  icu24 <- d$units %>% filter(time_from_admit_hours <= 24) %>%
    group_by(admission_id) %>% summarize(icu24 = as.integer(any(unit_type=="ICU")), .groups="drop")
  idv <- idv %>% left_join(icu24, by="admission_id") %>% mutate(icu24 = ifelse(is.na(icu24),0,icu24))
  
  # Urgent/emergent vs elective
  idv$urgent <- as.integer(cohort$admission_type %in% c("Urgent","Emergency"))
  
  # Charlson (prior 1 year)
  dx_prior <- d$dx %>%
    mutate(days_from_admit = as.numeric(difftime(dx_date, cohort$admit_time[match(admission_id, cohort$id)], units="days"))) %>%
    filter(days_from_admit <= 0 & days_from_admit >= -365)
  cci <- comorbidity::comorbidity(
    x = dx_prior, id="admission_id", code="icd10_code", map="charlson_icd10_quan",
    assign0 = TRUE, tidy.codes = TRUE
  ) %>% as_tibble()
  cci$charlson_sum <- rowSums(cci %>% select(-admission_id))
  idv <- idv %>% left_join(cci %>% select(admission_id, charlson_sum), by="admission_id") %>%
    mutate(charlson_sum = replace_na(charlson_sum, 0L))
  
  # Medications within 24h: UFH/LMWH, sedatives/hypnotics, anticholinergics, antipsychotics, steroids
  meds24 <- d$meds %>%
    filter(start_time <= (cohort$admit_time[match(admission_id, cohort$id)] + lubridate::hours(24))) %>%
    mutate(med_class = case_when(
      str_detect(tolower(med_name), "heparin|enoxaparin|dalteparin|fondaparinux") ~ "parenteral_anticoag",
      str_detect(tolower(med_name), "midazolam|lorazepam|zolpidem|eszopiclone|triazolam") ~ "sed_hyp",
      str_detect(tolower(med_name), "diphenhydramine|oxybutynin|amitriptyline|scopolamine|benztropine") ~ "anticholinergic",
      str_detect(tolower(med_name), "haloperidol|olanzapine|quetiapine|risperidone|ziprasidone") ~ "antipsychotic",
      str_detect(tolower(med_name), "predni|methylpred|dexamethasone|hydrocortisone") ~ "steroid",
      TRUE ~ NA_character_
    )) %>% filter(!is.na(med_class)) %>% distinct(admission_id, med_class) %>%
    pivot_wider(names_from=med_class, values_from=med_class, values_fn = length, values_fill = 0) %>%
    mutate(across(everything(), ~ as.integer(.>0)))
  idv <- idv %>% left_join(meds24, by="admission_id") %>%
    mutate(across(c(parenteral_anticoag, sed_hyp, anticholinergic, antipsychotic, steroid), ~replace_na(.,0L)))
  
  # Cognition: CAM-ICU or confusion on admission
  cog <- d$nursing %>% filter(assessment_time <= (cohort$admit_time[match(admission_id, cohort$id)] + lubridate::hours(24))) %>%
    group_by(admission_id) %>% summarize(delirium_on_admit = as.integer(any(cam_icu_positive==1, na.rm=TRUE)), .groups="drop")
  idv <- idv %>% left_join(cog, by="admission_id") %>% mutate(delirium_on_admit = replace_na(delirium_on_admit,0L))
  
  # Baseline function/mobility within 24h
  fn <- d$nursing %>% filter(assessment_time <= (cohort$admit_time[match(admission_id, cohort$id)] + lubridate::hours(24))) %>%
    select(admission_id, ampac_mobility_raw, barthel_total) %>%
    group_by(admission_id) %>% summarize(
      ampac = suppressWarnings(max(ampac_mobility_raw, na.rm=TRUE)),
      barthel = suppressWarnings(max(barthel_total, na.rm=TRUE)),
      .groups="drop"
    )
  fn$ampac[is.infinite(fn$ampac)] <- NA
  fn$barthel[is.infinite(fn$barthel)] <- NA
  # Standardization: rescale to 0-100 then z-scored over analytic set later
  idv <- idv %>% left_join(fn, by="admission_id")
  
  idv
}

# Outcomes
derive_outcomes <- function(d, cohort){
  out <- cohort %>% select(admission_id = id) %>% mutate(
    los_days = pmax(1, ceiling(cohort$los_hours/24))
  )
  # Deconditioning (decline in AM-PAC >=3 or Barthel >=10)
  fn_all <- d$nursing %>% arrange(admission_id, assessment_time) %>%
    group_by(admission_id) %>% summarize(
      ampac_first = dplyr::first(ampac_mobility_raw),
      ampac_last  = dplyr::last(ampac_mobility_raw),
      barthel_first = dplyr::first(barthel_total),
      barthel_last  = dplyr::last(barthel_total),
      .groups="drop"
    )
  out <- out %>% left_join(fn_all, by="admission_id") %>%
    mutate(
      dec_ampac = (ampac_first - ampac_last) >= 3,
      dec_barthel = (barthel_first - barthel_last) >= 10,
      deconditioning = as.integer(dplyr::coalesce(dec_ampac, FALSE) | (!isTRUE(dec_ampac) & dplyr::coalesce(dec_barthel, FALSE)))
    )
  # VTE outcome: ICD plus imaging window [-1,+3] days
  vte_dx <- d$dx %>% filter(grepl(icd10_pe_pattern, icd10_code) | grepl(icd10_dvt_pattern, icd10_code)) %>%
    select(admission_id, icd10_code, dx_date)
  img <- d$imaging %>% filter(cpt_code %in% c(cpt_ctpa, cpt_venous_duplex)) %>% select(admission_id, cpt_code, order_time)
  vte_flag <- vte_dx %>% left_join(img, by="admission_id") %>%
    mutate(delta = as.numeric(difftime(order_time, dx_date, units="days"))) %>%
    group_by(admission_id) %>% summarize(vte = as.integer(any(delta >= -1 & delta <= 3, na.rm=TRUE)), .groups="drop")
  out <- out %>% left_join(vte_flag, by="admission_id") %>% mutate(vte = replace_na(vte,0L))
  
  # Falls from safety system
  falls <- d$safety %>% filter(tolower(event_type)=="fall") %>% distinct(admission_id) %>% mutate(fall=1L)
  out <- out %>% left_join(falls, by="admission_id") %>% mutate(fall = replace_na(fall,0L))
  
  # Pressure injuries stage >=2: derive from dx codes
  pi <- d$dx %>% filter(grepl(icd10_pressure_ulcer_pattern, icd10_code)) %>%
    mutate(stage2plus = grepl("2|3|4|6", icd10_code)) %>% # 6 used for deep tissue damage (L89.x6)
    group_by(admission_id) %>% summarize(pressure_ulcer = as.integer(any(stage2plus, na.rm=TRUE)), .groups="drop")
  out <- out %>% left_join(pi, by="admission_id") %>% mutate(pressure_ulcer = replace_na(pressure_ulcer,0L))
  
  # Prolonged LOS >7 days
  out$prolonged_los <- as.integer(out$los_days > 7)
  
  # Disposition: post-acute facility vs home
  out$dispo_postacute <- as.integer(cohort$discharge_disposition %in% c(dispo_post_acute))
  
  # 30-day readmission (requires linkage across facilities); placeholder flag column to be joined by caller
  out$readmit30 <- 0L
  out
}
```
