## Supplementary material for "Validation of the Immobility Harm Risk Score for Predicting Inpatient Immobility-Associated Outcomes: A Retrospective Cohort Study": R code: 04_impute.R

```
# 04_impute.R — multiple imputation
impute_predictors <- function(df){
  library(mice)
  # Rescale AM-PAC (6–24) and Barthel (0–100) to 0–100 for harmonization
  df$ampac_scaled <- ifelse(!is.na(df$ampac), (df$ampac - 6) / (24 - 6) * 100, NA_real_)
  df$barthel_scaled <- ifelse(!is.na(df$barthel), df$barthel, NA_real_)
  df$baseline_mobility <- dplyr::coalesce(df$ampac_scaled, df$barthel_scaled)
  
  keep <- df %>% select(age, sex, race_ethnicity, principal_icd10, icu24, urgent,
                        charlson_sum, parenteral_anticoag, sed_hyp, anticholinergic, antipsychotic, steroid,
                        delirium_on_admit, baseline_mobility, ehr)
  # Simple encodings
  keep$sex <- factor(keep$sex)
  keep$race_ethnicity <- factor(keep$race_ethnicity)
  keep$icu24 <- factor(keep$icu24)
  keep$urgent <- factor(keep$urgent)
  keep$ehr <- factor(keep$ehr)
  
  meth <- make.method(keep)
  pred <- make.predictorMatrix(keep)
  meth["age"] <- "pmm"
  meth["charlson_sum"] <- "pmm"
  meth["baseline_mobility"] <- "pmm"
  meth[c("sex","race_ethnicity","icu24","urgent","ehr",
         "parenteral_anticoag","sed_hyp","anticholinergic","antipsychotic","steroid",
         "delirium_on_admit")] <- "logreg"
  
  imp <- mice(keep, m=10, maxit=20, method=meth, predictorMatrix=pred, seed=42, printFlag=FALSE)
  imp
}
```
