## Supplementary material for "Validation of the Immobility Harm Risk Score for Predicting Inpatient Immobility-Associated Outcomes: A Retrospective Cohort Study": R code: 05_model_fit.R

```
# 05_model_fit.R — elastic net logistic for 7 outcomes
fit_models <- function(imp, X, Y_list, alpha=0.5){
  library(glmnet)
  fits <- list()
  for (nm in names(Y_list)){
    y <- Y_list[[nm]]
    # glmnet expects matrix
    xmat <- model.matrix(~ . , data=X)[,-1]
    # Class weights for imbalance
    w <- ifelse(y==1, 1/mean(y), 1/(1-mean(y)))
    cv <- cv.glmnet(xmat, y, family="binomial", alpha=alpha, weights=w, nfolds=10, type.measure="deviance")
    fit <- glmnet(xmat, y, family="binomial", alpha=alpha, lambda=cv$lambda.1se)
    fits[[nm]] <- list(fit=fit, lambda=cv$lambda.1se)
  }
  fits
}

predict_suite <- function(fits, newdata){
  xmat <- model.matrix(~ . , data=newdata)[,-1]
  sapply(fits, function(obj) as.numeric(predict(obj$fit, newx=xmat, type="response")))
}
```
