## Supplementary material for "Validation of the Immobility Harm Risk Score for Predicting Inpatient Immobility-Associated Outcomes: A Retrospective Cohort Study": R code: 06_validate.R

```
# 06_validate.R — internal CV, bootstrap, temporal validation, calibration, DCA, NRI
library(pROC); library(ResourceSelection); library(rmda); library(nricens)

auc_ci <- function(y, p){
  rocobj <- pROC::roc(y, p, quiet=TRUE)
  ci <- pROC::ci.auc(rocobj)
  c(auc=as.numeric(rocobj$auc), lwr=ci[1], upr=ci[3])
}

brier <- function(y,p) mean((p - y)^2)

calibration <- function(y,p){
  mod <- glm(y ~ qlogis(p), family=binomial())
  c(int = coef(mod)[1], slope = coef(mod)[2])
}

hoslem <- function(y,p,g=10){
  ResourceSelection::hoslem.test(y, p, g=g)$p.value
}

decision_curve_util <- function(y,p, thresholds = seq(0.05,0.20,by=0.01)){
  df <- data.frame(y=y, p=p)
  dca(p ~ 1, data=df, thresholds=thresholds, confidence.intervals=FALSE)
}

nri_cf <- function(y, p_new, p_old){
  nricens::nribin(event = y, p.std = p_old, p.new = p_new, updown = 'category-less')
}
```
