## Supplementary material for "Validation of the Immobility Harm Risk Score for Predicting Inpatient Immobility-Associated Outcomes: A Retrospective Cohort Study": R code: 07_comparators.R

```
# 07_comparators.R — Braden, Morse, Padua, Charlson, LACE, etc.

compute_braden <- function(nursing_row){
  # Assumes total already supplied in EHR (6-23). Return as-is.
  nursing_row$braden_total
}

compute_morse <- function(nursing_row){
  nursing_row$morse_total
}

compute_padua <- function(row, dx_prior_codes){
  score <- 0
  add <- function(points) score <<- score + points
  # Operate on row (age, bmi, med flags) + dx_prior_codes vector
  if (!is.na(row$active_cancer) && row$active_cancer==1) add(3)
  if (!is.na(row$prior_vte) && row$prior_vte==1) add(3)
  if (!is.na(row$reduced_mobility) && row$reduced_mobility==1) add(3)
  if (!is.na(row$thrombophilia) && row$thrombophilia==1) add(3)
  if (!is.na(row$recent_trauma_surg) && row$recent_trauma_surg==1) add(2)
  if (!is.na(row$age) && row$age >= 70) add(1)
  if (!is.na(row$hf_or_rf) && row$hf_or_rf==1) add(1)
  if (!is.na(row$acute_mi_or_stroke) && row$acute_mi_or_stroke==1) add(1)
  if (!is.na(row$infection_rheum) && row$infection_rheum==1) add(1)
  if (!is.na(row$bmi) && row$bmi >= 30) add(1)
  if (!is.na(row$hormonal_tx) && row$hormonal_tx==1) add(1)
  score
}

compute_lace <- function(los_days, urgent, charlson_sum, ed_visits_6mo){
  # Teh et al. and Staples et al. supplementary table
  L <- dplyr::case_when(
    los_days == 0 ~ 0,
    los_days == 1 ~ 1,
    los_days == 2 ~ 2,
    los_days == 3 ~ 3,
    los_days >= 4 & los_days <= 6 ~ 4,
    los_days >= 7 & los_days <= 13 ~ 5,
    los_days >= 14 ~ 7,
    TRUE ~ 0
  )
  A <- ifelse(urgent==1, 3, 0)
  C <- dplyr::case_when(
    charlson_sum == 0 ~ 0,
    charlson_sum == 1 ~ 1,
    charlson_sum == 2 ~ 2,
    charlson_sum >= 3 ~ 3
  )
  E <- dplyr::case_when(
    ed_visits_6mo == 0 ~ 0,
    ed_visits_6mo == 1 ~ 1,
    ed_visits_6mo == 2 ~ 2,
    ed_visits_6mo >= 3 ~ 3
  )
  as.integer(L + A + C + E)
}
```
