## Supplementary material for "Validation of the Immobility Harm Risk Score for Predicting Inpatient Immobility-Associated Outcomes: A Retrospective Cohort Study": R code: 08_metrics_plots.R

```
# 08_metrics_plots.R — plots for calibration, decision curves, subgroup forest
library(ggplot2)

plot_calibration <- function(y, p){
  df <- data.frame(y=y, p=p) %>%
    mutate(bin = ntile(p, 10)) %>%
    group_by(bin) %>% summarize(obs = mean(y), pred = mean(p)) %>% ungroup()
  ggplot(df, aes(x=pred, y=obs)) +
    geom_point() + geom_line() +
    geom_abline(slope=1, intercept=0, linetype="dashed") +
    labs(x="Predicted risk", y="Observed risk", title="Calibration plot (deciles + 45°)") +
    theme_minimal()
}

plot_dca <- function(y, p){
  thresholds <- seq(0.05,0.20,by=0.01)
  nb <- sapply(thresholds, function(t){ mean( (p>t)*y - (p>t)*(1-y)*t/(1-t) ) })
  df <- data.frame(threshold=thresholds, net_benefit = nb)
  ggplot(df, aes(threshold, net_benefit)) + geom_line() +
    labs(x="Threshold probability", y="Net benefit", title="Decision Curve") +
    theme_minimal()
}

plot_forest_subgroup_auc <- function(res_by_group){
  # res_by_group: data.frame(group, auc, lwr, upr)
  ggplot(res_by_group, aes(x=group, y=auc, ymin=lwr, ymax=upr)) +
    geom_pointrange() + coord_flip() + theme_minimal() +
    labs(y="AUC", x="Subgroup", title="Subgroup discrimination")
}
```
