## Supplementary material for "Validation of the Immobility Harm Risk Score for Predicting Inpatient Immobility-Associated Outcomes: A Retrospective Cohort Study": R code: 09_tables_docx.R

```
# 09_tables_docx.R — build Word doc with tables using officer/flextable
library(officer); library(flextable); library(dplyr); library(stringr)

make_docx <- function(output = "medrxiv_supplement.docx"){
  doc <- read_docx()
  doc <- body_add_par(doc, "Supplementary Materials", style = "heading 1")
  
  # Table S1 — Codes
  doc <- body_add_par(doc, "Table S1. Code lists for outcomes and dispositions", style="heading 2")
  tbl1 <- tibble::tibble(
    Domain = c("Pulmonary embolism (ICD-10-CM)", "DVT (ICD-10-CM)", "Pressure injury stage ≥2 (ICD-10-CM)", "CTPA (CPT)", "LE venous duplex (CPT)", "Discharge to post-acute (NUBC)"),
    Codes = c("I26.* (all 4th/5th digit expansions including I26.90, I26.92–I26.95); exclude chronic I27.82 when not acute",
              "I82.4*, I82.5*, I82.9* (lower-extremity deep veins families)",
              "L89.* with stage 2–4 or deep tissue damage (x=2–4 or 6)",
              "71275",
              "93970 (bilateral complete), 93971 (unilateral/limited)",
              "03 (SNF), 61 (Swing bed), 62 (IRF), 63 (LTCH), 64 (Medicaid NF)")
  )
  doc <- body_add_flextable(doc, flextable(tbl1))
  
  # Table S2 — AM-PAC & Barthel mapping to standardized z
  doc <- body_add_par(doc, "Table S2. Mapping AM-PAC ‘6-Clicks’ and Barthel to standardized z-scores", style="heading 2")
  desc <- "Algorithm: (1) Rescale AM-PAC Basic Mobility raw 6–24 to 0–100 using (raw-6)/18*100; use Barthel 0–100 as-is. (2) Harmonized Mobility = coalesce(AM-PAC_rescaled, Barthel). (3) Standardize within analytic sample: z = (Harmonized Mobility − mean)/sd. Example grid below uses illustrative mean=55, sd=15."
  grid <- tibble::tibble(
    Instrument = c("AM-PAC", "AM-PAC", "Barthel", "Barthel"),
    Raw = c(12, 20, 40, 85),
    Rescaled_0_100 = c((12-6)/18*100, (20-6)/18*100, 40, 85),
    z_if_mean55_sd15 = round((c((12-6)/18*100,(20-6)/18*100,40,85)-55)/15,2)
  )
  doc <- body_add_par(doc, desc, style="Normal")
  doc <- body_add_flextable(doc, flextable(grid))
  
  # Table S3 — CMS Planned Readmission Exclusions (summary)
  doc <- body_add_par(doc, "Table S3. CMS Planned Readmission Algorithm v4.0 categories", style="heading 2")
  cms <- tibble::tibble(
    Category = cms_planned_categories,
    Notes = c("Delivery-related readmissions", "Transplant aftercare and maintenance", "Chemotherapy maintenance",
              "Radiation therapy maintenance", "Inpatient rehabilitation", "Elective major procedures (pre-specified lists)")
  )
  doc <- body_add_flextable(doc, flextable(cms))
  
  # Table S7 — Padua operationalization
  doc <- body_add_par(doc, "Table S7. Padua variables and EHR operationalization", style="heading 2")
  doc <- body_add_flextable(doc, flextable(padua_vars))
  
  # Text S1 — Session info (template)
  doc <- body_add_par(doc, "Text S1. R Session Info", style="heading 2")
  sess <- paste0(capture.output(sessionInfo()), collapse="\n")
  doc <- body_add_par(doc, sess, style="Normal")
  
  # Text S2 — Minimal scoring function
  doc <- body_add_par(doc, "Text S2. Minimal scoring function (R)", style="heading 2")
  code <- '
score_bedbike <- function(row, coefs){
  # row: named numeric vector of predictors matching model matrix columns
  # coefs: named list of numeric coefficient vectors per outcome (first element intercept)
  probs <- vapply(names(coefs), function(outc){
    b <- coefs[[outc]]
    xb <- b[1] + sum(row * b[-1], na.rm=TRUE)
    1/(1+exp(-xb))
  }, numeric(1))
  return(probs)
}
'
  doc <- body_add_par(doc, code, style="Normal")
  
  # Text S3 — TRIPOD checklist pointer (narrative)
  doc <- body_add_par(doc, "Text S3. TRIPOD items (narrative compliance)", style="heading 2")
  tripod <- "We report: source of data; participants (eligibility, setting, dates); predictors (definitions, timing, blinding); outcomes (definitions, windows); sample size; missing data (MICE); model specification (elastic net logistic with α=0.5); internal validation (10-fold CV + bootstrap); temporal validation (2013–2019 train; 2020–2023 test); performance (AUC, Brier, calibration intercept/slope, HL test); clinical utility (DCA); reclassification (category-free NRI); subgroup analyses; availability of code and code lists; PROBAST appraisal. A completed itemized TRIPOD table is included in the manuscript main Supplement."
  doc <- body_add_par(doc, tripod, style="Normal")
  
  print(doc, target = output)
  return(output)
}
```
