## Supplementary material for "Validation of the Immobility Harm Risk Score for Predicting Inpatient Immobility-Associated Outcomes: A Retrospective Cohort Study": R code: 10_score_function.R

```
# 10_score_function.R — exported scoring from fitted coefficients
# This file expects a saved RDS of coefficients per outcome created by 05_model_fit.R
scorebedbike <- function(newdata, coefs_path="artifacts/coef_list.rds"){
  coefs <- readRDS(coefs_path)
  xmat <- model.matrix(~ ., data=newdata)[,-1]
  probs <- sapply(coefs, function(b){
    as.numeric(1/(1+exp(-(b[1] + xmat %*% b[-1]))))
  })
  as_tibble(probs)
}
```
