## Supplementary material for "Validation of the Immobility Harm Risk Score for Predicting Inpatient Immobility-Associated Outcomes: A Retrospective Cohort Study": R code: run_all.R

```
# run_all.R — orchestrates pipeline (expects CSVs under data/)
source("R/00_setup.R")
source("R/01_data_spec.R")
source("R/02_code_lists.R")
source("R/03_extract_transform.R")
source("R/04_impute.R")
source("R/05_model_fit.R")
source("R/06_validate.R")
source("R/07_comparators.R")
source("R/08_metrics_plots.R")
source("R/09_tables_docx.R")
source("R/10_score_function.R")

dir.create("artifacts", showWarnings = FALSE)

d <- read_all("data")

cohort <- construct_cohort(d)
Xraw <- derive_predictors(d, cohort)
Y <- derive_outcomes(d, cohort)

# Exclusions for missing key predictors (>20% missing across key set already handled at data entry)
# Impute
imp <- impute_predictors(Xraw)

# Complete model matrix from first imputed set for fitting (for simplicity)
Xcomp <- complete(imp, action=1)
Y_list <- list(
  deconditioning = Y$deconditioning,
  vte = Y$vte,
  fall = Y$fall,
  pressure_ulcer = Y$pressure_ulcer,
  prolonged_los = Y$prolonged_los,
  dispo_postacute = Y$dispo_postacute,
  readmit30 = Y$readmit30
)

fits <- fit_models(imp, Xcomp, Y_list, alpha=0.5)

# Save coefficients
coef_list <- lapply(fits, function(obj){
  b <- as.numeric(coef(obj$fit))
  names(b) <- rownames(coef(obj$fit))
  b
})
saveRDS(coef_list, file="artifacts/coef_list.rds")

# Metrics (apparent; for full CV/bootstrapping see 06_validate.R — customize as needed)
probs <- predict_suite(fits, Xcomp) %>% as_tibble()
metrics <- tibble::tibble(
  outcome = names(Y_list),
  auc = mapply(function(y, p) auc_ci(y, p)[1], Y_list, probs),
  brier = mapply(brier, Y_list, probs),
  hl_p = mapply(hoslem, Y_list, probs)
)
readr::write_csv(metrics, "artifacts/metrics_apparent.csv")

# Figures — create example placeholders from apparent predictions
dir.create("artifacts/figures", showWarnings = FALSE)
for (nm in names(Y_list)){
  png(file.path("artifacts/figures", paste0("calibration_", nm, ".png")), width=1200, height=900, res=150)
  print(plot_calibration(Y_list[[nm]], probs[[nm]]))
  dev.off()
  
  png(file.path("artifacts/figures", paste0("dca_", nm, ".png")), width=1200, height=900, res=150)
  print(plot_dca(Y_list[[nm]], probs[[nm]]))
  dev.off()
}

# Build Supplement docx with static tables that do not require patient-level data.
make_docx(output = "artifacts/medrxiv_supplement.docx")

message("Pipeline complete. See artifacts/ for outputs.")
```
